# National and regional prevalence of SARS-CoV-2 antibodies in primary and secondary school children in England: the School Infection Survey, a national open cohort study, November 2021

**DOI:** 10.1101/2022.07.20.22277838

**Authors:** Annabel A Powell, Georgina Ireland, Rebecca Leeson, Andrea Lacey, Ben Ford, John Poh, Samreen Ijaz, Justin Shute, Peter Cherepanov, Richard Tedder, Christian Bottomley, Fiona Dawe, Punam Mangtani, Peter Jones, Patrick Nguipdop-Djomo, Shamez Ladhani

## Abstract

**Background:** Risk factors for infection and, therefore, antibody positivity rates will be different in children compared to adults. We aim to estimate national and regional prevalence of SARS-CoV-2 antibodies in primary (4-11-year-olds) and secondary (11-15-year-olds) school children between 10 November and 10 December 2021.

**Methods:** Cross-sectional surveillance in England using two stage sampling, firstly stratifying into regions and selecting local authorities, then selecting schools according to a stratified sample within selected local authorities. Participants were sampled using a novel oral fluid validated assay for SARS-CoV-2 spike and nucleocapsid IgG antibodies.

**Results:** 4,980 students from 117 state-funded schools (2,706 from 83 primary schools, 2,274 from 34 secondary schools) provided a valid sample. After weighting for age, sex and ethnicity, and adjusting for assay accuracy, the national prevalence of SARS-CoV-2 antibodies in primary school students, who were all unvaccinated, was 40.1% (95%CI; 37.3-43.0). Antibody prevalence increased with age (p<0.001) and were higher in urban than rural schools (p=0.01). In secondary school students, the adjusted, weighted national prevalence of SARS-CoV-2 antibodies was 82.4% (95%CI; 79.5-85.1); including 57.5% (95%CI; 53.9-61.1) in unvaccinated and 97.5% (95%CI; 96.1-98.5) in vaccinated students. Antibody prevalence increased with age (p<0.001), and was not significantly different in urban versus rural students (p=0.1).

**Conclusions:** Using a validated oral fluid assay, we estimated national and regional seroprevalence of SARS-CoV-2 antibodies in primary and secondary school students. In November 2021, 40% of primary school students and nearly all secondary school students in England had SARS-CoV2 antibodies through a combination of natural infection and vaccination.

## Introduction

Children have a very low risk of severe or fatal COVID-19 compared to adults and typically remain asymptomatic or develop mild, transient upper respiratory tract symptoms when infected with SARS-CoV-2, the virus responsible for COVID-19.(1) Early pandemic, investigations using PCR-testing in symptomatic individuals suggested that children may be less susceptible to SARS-CoV-2 infection,(1) but subsequent studies, importantly using SARS-CoV-2 antibodies to measure both symptomatic and asymptomatic infections, indicated a bigger role for children in infection and transmission of the virus.(2) Additionally, we and others have shown that children develop robust humoral and cellular immune responses against SARS-CoV-2 irrespective of symptomatic or asymptomatic infection.(3)

In the UK, the first COVID-19 vaccines were authorised in December 2020 and offered to those at highest risk, starting with older adults, healthcare workers and those with comorbidities, followed by progressively younger adults, with 16-17-year-olds offered the vaccine from 4 August 2021.(4) Vaccination for high-risk 12-15-year-olds began on 31 August 2021,(5) but, on 3 September 2021, the UK Joint Committee on Vaccination and Immunisation (JCVI) recommended against vaccinating healthy 12-15-year-olds because of marginal risk-benefits and concerns about the risk of vaccine-induced myocarditis in young men.(5) This decision was, however, overturned by UK ministers on 14 September 2021 and the vaccine was subsequently offered to this age-group to reduce educational disruption.(6) In 5-11-year-olds, a lower dose of Pfizer-BioNTech vaccine (10 μg/dose) was offered to high-risk children from February 2022, with a plan to offer the vaccine to all 5-11-year-olds from April 2022. (7)

The UK has been proactive in keeping schools open throughout the pandemic, ensuring that they were the last settings to close and first to reopen. All schools were closed following the first national lockdown in March 2020, and then partially reopening for some academic years in June 2020 with extensive mitigations in place.(8) In-school studies showed limited infection and transmission among staff or students, which facilitated the full reopening of all school years for in-person learning from September 2020.(9) In school-aged children, case rates increased during September-October 2020, with further increases in December 2020 with the emergence of the Alpha variant, during June-July 2021 because of the Delta variant, and during September-December 2021, when all mitigations were removed from schools. (10-12)

As part of national surveillance, the UK Health Security Agency (UKHSA) developed and validated an oral fluid (OF) assay to facilitate large-scale community SARS-CoV-2 seroprevalence studies.(13) This assay was initially used in a large national Schools Infection Survey (SIS-1), an open cohort study of SARS-CoV-2 infection and transmission in state-funded educational settings during the 2020/21 academic year.(14) For the 2021/22 academic year (SIS-2), the methodology was revised and extended to include a nationally- and regionally-representative sample of primary and secondary school-aged children.(15) Additionally, the OF assay was augmented to distinguish between infection- and vaccine-induced antibodies. Here, we report the first results of SIS2 with national and regional prevalence of SARS-CoV-2 antibodies in primary and secondary-school aged children during November/December 2021 in England.

## Methods

### Sampling Methodology

The schools were identified from a list of state-funded schools held by the Department for Education (DfE) and were equally distributed across English regions. Independent schools, special education schools, pupil referral units, further education colleges and home-schooled children were excluded. Only sixth forms attached to or part of secondary schools were included. Of note, 47 schools that had participated in SIS1 also participated in SIS2 and formed part of the selected sample.

In participating schools, parents were invited to enrol their children on a voluntary basis. An online consent and survey form were sent to all parents/students using the Smart Survey platform. Detailed methodology of the sampling frame and approach can be found online.(15) Parents of children from Reception to Year 11 (4-16-year-olds) gave consent and enrolled on behalf of their children, while students in Years 12 and 13 (16-18-year-olds) consented and enrolled themselves. Trained Office for National Statistics (ONS) field staff attended participating schools on pre-specified testing days and provided written and verbal instruction on self-sampling for SARS-CoV-2 antibodies using the Oracol OF collection device (Malvern Medical Developments Ltd, UK). The samples were processed and tested by UKHSA.

### Laboratory testing

OF samples were processed and tested for antibodies as described previously.(13) Briefly, OF was extracted by adding the elution buffer and agitating the swab. The resulting eluate was centrifuged, collected, and stored at -30°C until testing. OF samples were tested for SARS-CoV-2 antibodies using an in-house, Immunoglobulin G antibody capture enzyme immunoassay (EIA) based on a solid-phase anti-human IgG with either a HRP-conjugated Nucleoprotein (N) antigen probe,(13) or a HRP-conjugated Spike (S) S1 subunit protein probe. The SARS-CoV-2 S1 antigen was produced and gifted by The Francis Crick Institute.(16) In unvaccinated children and adults, the NP assay had a sensitivity and specificity of 80 % and 99%, respectively, for the NP assay,(13) and 75% and 98%, respectively, for the S1 assay (unpublished data).

### Sample Size and sampling design

The original SIS-1 study had oversampled schools in high SARS-CoV-2 prevalence areas in September 2020 to understand virus transmission risks and dynamics in educational settings.(14) Sampled schools were expanded for SIS2, to produce robust estimates of SARS-CoV-2 antibody prevalence in school children at a regional and national level. We calculated the sample size for the most statistically conservative scenario of 50% antibodies prevalence, which would result in the widest confidence intervals, and aimed for ±5% precision with 95% confidence at a regional level for primary and secondary schools. Assumptions on school’s average population (280 students in primary, 965 in secondary schools) and school-level response rate (15%) were based on administrative data and SIS-1, respectively. After accounting for clustering within schools (design effect 2.3 based on SIS-1), the estimated sample size was 13 primary and 7 secondary schools in each of the nine regions, for a total of 180 schools nationally.

Schools were selected using a two-stage self-weighted sampling scheme, stratified by region and primary versus secondary schools, as used for SIS1. The primary sampling unit (PSU) in each region was the local authority area (LA). LAs were sorted using the percentage of primary schools located in urban areas within the LA, to ensure sampled LAs were representative of the urban/rural split in the region, and five LAs were selected with probability proportional to size (number of primary schools) to increase likelihood of sampling LAs with more schools. For practical purposes, given that SIS-2 was expanding on SIS-1, LAs already included in SIS-1 but not selected in the random sample above were included in replacement of an LA with a similar percentage of primary schools and total number of schools.

The second sampling unit was the school, stratified by regions and by primary versus secondary schools, and selected using systematic random sampling. Schools’ sampling was implicitly stratified on percentage of students eligible for free school meals (FSM – a proxy-measure of socio-economic deprivation) and presence of Sixth Form (16-18-year-olds) for secondary schools), by sorting on these criteria prior to sampling. All students in selected schools were eligible to participate. Similar to the LA-level, schools already participating in SIS1 replaced any newly sampled schools with similar characteristics (**Supplement Table 1**).

### Statistical analysis

We calculated antibody prevalence overall and by groups, using the number of participants with antibody levels above the detection threshold as the numerator, and the total number of participants with a valid result as the denominator. Those with inconclusive or no results (e.g. missing or poor sample quality) were excluded. Prevalence estimates were adjusted for diagnostic test sensitivity and specificity in unvaccinated participants as detailed above. All estimates were weighted to be representative at regional and national levels, accounting for sampling design, age, sex and ethnicity. The 95%CI were calculated using robust standard error to account for the cluster sampling design.

Direct deterministic linkage using personal identifying information was used to link participant data to national laboratory reports of SARS-CoV-2 PCR and LFD tests (Second Generation Surveillance System, SGSS) to ascertain prior SARS-CoV-2 infection and to the National Immunisation Management Service (NIMS) database to identify COVID-19 vaccination status and date of vaccination, both held at UKHSA. As older students had only just become eligible for their second dose in November 2021, students were grouped as unvaccinated or vaccinated if they had received ≥1 COVID-19 vaccine dose ≥14 days before their antibody test. SARS-CoV-2 antibody prevalence are reported for primary and secondary school-aged students nationally, by region and by vaccination status. In unvaccinated children, the presence of N-antibody and/or S-antibody was considered as evidence of past infection; antibody prevalence was adjusted to reflect the sensitivity and specificity of the OF assay compared to serum. In vaccinated participants, no adjustments were made because of the very high antibody levels achieved by mRNA vaccines and the consequent high correlation between serum and OF antibody levels (data not shown). Non-overlapping 95%CI were used to assess statistically significant differences between groups. Where shown, P-values were estimated using the Chi-squared test.

For comparison with community SARS-CoV-2 antibody seroprevalence in young adults, regional seroprevalence data from NHS Blood and Transplant (NHS BT) serosurveillance of blood donors were obtained for 17-30-year-olds from 01 November to 03 December 2021. Donor sera are tested for N- and S-antibodies on Roche Elecsys Anti-SARSCoV-2 N assay and seroprevalence results are weighted using age, sex, and region. Data were managed in Microsoft Access and analysed in Stata SE (version 15.1).

## Results

In this first round of testing (10 November to 10 December 2021), 183 primary and 125 secondary schools were approached to participate (**Figure 1**). Overall, 117 schools (83 primary, 34 secondary) enrolled, and 5,972 students (3,183 primary, 2,789 secondary) were eligible for testing on the day and, after excluding 313 samples which were void, 4,980 results were included (2,706 primary, 2,274 secondary). Participant response rate was 10% (15% primary, 8% secondary). Participants were broadly similar by sex in primary and secondary schools, mostly from White ethnic background (86.1% primary, 89.3% secondary), and in urban locations (80.6% primary, 88.7% secondary) (**Table 1**). Linkage with SGSS showed that 13.3% (359/2,706) of primary and 19.5% (443/2,276) of secondary school students had ≥1 confirmed SARS-CoV-2 infection >2 weeks prior to OF sampling.

**Table 1.** **Characteristics of primary and secondary school students in England participating in Round 1 (10 November – 10 December 2021) oral fluid sampling of SIS2**

|  | SIS2 Cohort |  |  |  |  |  | National Data* |  |  |
| --- | --- | --- | --- | --- | --- | --- | --- | --- | --- |
|  | Total |  | Primary |  | Secondary |  | Total | Primary | Secondary |
|  | n | % | n | % | n | % | % | % | % |
| Sex |  |  |  |  |  |  |  |  |  |
| Female | 2503 | 50.3 | 1337 | 49.4 | 1166 | 51.3 | 49.4 | 49.1 | 49.8 |
| Male | 2447 | 49.7 | 1369 | 50.1 | 1108 | 48.7 | 50.6 | 50.9 | 50.2 |
| Age group |  |  |  |  |  |  |  |  |  |
| 4-7 | 1333 | 26.8 | 1333 | 49.3 | - | - | 31.8 | 56.6 | - |
| 8-11 | 1867 | 37.5 | 1367 | 50.5 | 500 | 22.0 | 32.6 | 43.4 | 18.7 |
| 12-15 | 1648 | 33.1 | - | - | 1648 | 72.5 | 30.2 | - | 69.1 |
| 16-17 | 118 | 2.4 | - | - | 118 | 5.2 | 5.4 | - | 12.3 |
| Missing | 14 | 0.3 | 6 | 0.2 | 8 | 0.4 | - | - | - |
| Ethnicity |  |  |  |  |  |  |  |  |  |
| White | 4361 | 87.6 | 2330 | 86.1 | 2031 | 89.3 | 72.4 | 72.8 | 71.7 |
| Black | 56 | 1.1 | 33 | 1.2 | 23 | 1.0 | 5.4 | 5.4 | 6.2 |
| Asian | 196 | 3.9 | 130 | 4.8 | 66 | 2.9 | 12.0 | 11.8 | 12.1 |
| Mixed | 312 | 6.3 | 182 | 6.7 | 130 | 5.7 | 6.4 | 6.6 | 6.0 |
| Other | 55 | 1.1 | 31 | 1.2 | 24 | 1.1 | 3.7 | 3.4 | 4.0 |
| Region |  |  |  |  |  |  |  |  |  |
| East Midlands | 386 | 7.8 | 225 | 8.3 | 161 | 7.1 | 4.6 | 4.8 | 4.6 |
| East of England | 563 | 11.3 | 382 | 14.1 | 181 | 8.0 | 13.2 | 14.0 | 12.9 |
| London | 511 | 10.3 | 366 | 13.5 | 145 | 6.4 | 9.8 | 10.3 | 9.9 |
| North East | 737 | 14.8 | 383 | 14.2 | 354 | 15.6 | 8.4 | 8.6 | 8.6 |
| North West | 666 | 13.4 | 418 | 15.4 | 248 | 10.9 | 11.0 | 11.1 | 11.3 |
| South East | 536 | 10.8 | 297 | 11.0 | 239 | 10.5 | 11.3 | 11.2 | 11.5 |
| South West | 707 | 14.2 | 254 | 9.4 | 453 | 19.9 | 16.3 | 15.4 | 16.0 |
| West Midlands | 201 | 4.0 | 134 | 5.0 | 67 | 2.9 | 16.4 | 15.6 | 15.8 |
| Yorkshire and the Humber | 673 | 13.4 | 247 | 9.1 | 426 | 18.7 | 9.2 | 9.0 | 9.4 |
| Location type |  |  |  |  |  |  |  |  |  |
| Urban | 4199 | 84.3 | 2182 | 80.6 | 2017 | 88.7 | 85.5 | 84.2 | 88.1 |
| Rural | 781 | 15.7 | 524 | 19.4 | 257 | 11.3 | 14.5 | 15.8 | 11.9 |
| Vaccination status |  |  |  |  |  |  |  |  |  |
| Unvaccinated | 3977 | 79.9 | 2706 | 100 | 1271 | 55.9 | - | - | - |
| Vaccinated** | 1003 | 20.1 | - | 0 | 1003 | 44.1 | - | - | 37.8 |
| Free school meals |  |  |  |  |  |  |  |  |  |
| Yes | 848 | 17.0 | 627 | 23.2 | 221 | 9.7 | 20.8 | 21.6 | 18.9 |
| No | 3927 | 78.9 | 1931 | 71.4 | 1996 | 87.8 | - | - | - |
| Don't know | 151 | 3.0 | 104 | 3.8 | 47 | 2.1 | - | - | - |
| Missing | 54 | 1.1 | 44 | 1.6 | 10 | 0.4 | - | - | - |
| **Total** | **4980** |  | **2706** |  | **2274** |  | N/A | N/A | N/A |

\* National data for each of the categories except for vaccination status was found online here: [Schools, pupils and their characteristics, Academic Year 2020/21 – Explore education statistics – GOV.UK \(explore-education-statistics.service.gov.uk\)](https://explore-education-statistics.service.gov.uk/). Data for vaccination status was found online here: [National flu and COVID-19 surveillance reports: 2021 to 2022 season - GOV.UK \(www.gov.uk\)](https://www.gov.uk/government/publications/national-flu-and-covid-19-surveillance-reports-2021-to-2022-season)
\*\* Defined as vaccinated if participant received $\geq 1$ COVID-19 vaccine dose $\geq 14$ days before sample date

**Figure 1.**
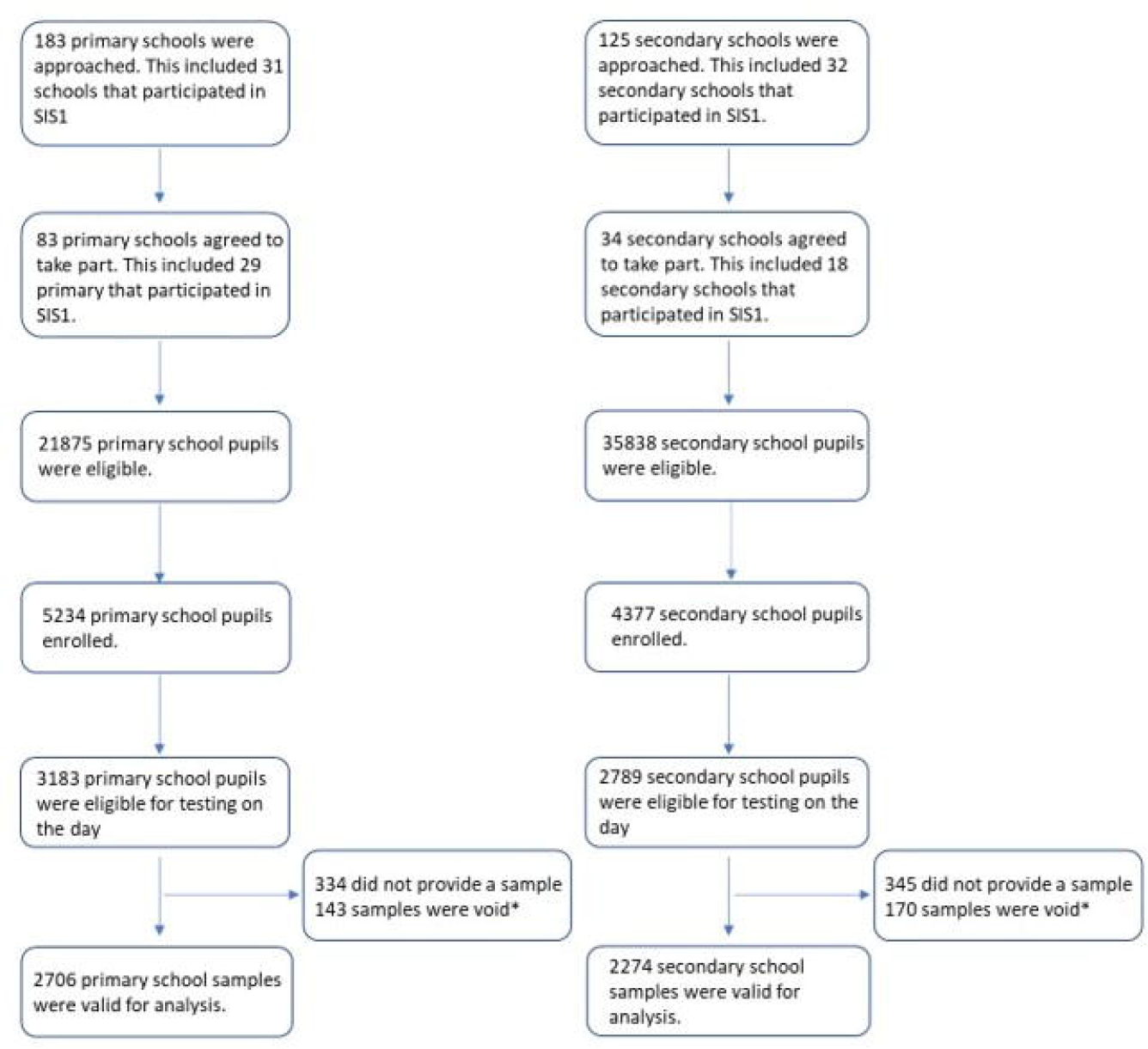
Flow chart of recruitment numbers achieved in SIS2 by school type and participants sampled. *Void tests are oral fluid samples that were rejected at the processing stage prior to antibody testing. They could be rejected based on number of reasons, most commonly where recovery of >2ml volume or

### Primary school students

Primary school students were unvaccinated and, therefore, antibody prevalence estimates reflect previous infection. Overall, 34.1% (924/2,706) of primary school students tested positive for SARS-CoV-2 antibodies. After weighting and adjusting for test sensitivity and specificity, national prevalence of SARS-CoV-2 antibodies in primary school-aged children was estimated at 40.1% (95%CI; 37.3-43.0%) (**Table 2**), regional estimates ranged from 21.9% (95%CI; 13.9-31.8) in the South West to 53.8% (95%CI; 43.1-64.3) in the West Midlands (**Table 3**). Antibody prevalence increased with age (p<0.001) (**Supplement Figure 2**), were higher in urban schools than rural (p=0.01) and there were no significant differences by sex (p=0.56).

**Table 2.** **Unweighted totals and weighted SARS-CoV-2 spike protein (S) and nucleocapsid (N) antibody positivity in unvaccinated primary school children and by COVID-19 vaccination status in secondary school children during Round 1 (10 November – 10 December 2021) in the SIS2 study**

|  | Primary |  | Secondary |  |  |  |  |  |
| --- | --- | --- | --- | --- | --- | --- | --- | --- |
|  | Total |  | Total |  | Vaccinated |  | Unvaccinated |  |
|  | n* | Weighted (%) ** | n* | Weighted (%) ** | n* | Weighted (%) ** | n* | Weighted (%) ** |
| Negative | 1782 | - | 620 | - | 35 | - | 585 | - |
| Total Positive | 924 | 32.7% (30.0-35.4) | 1654 | 75.2% (71.9-78.2) | 968 | 97.5% (96.1-98.5) | 686 | 57.5% (53.9-61.1) |
| S positive / N positive | 627 | 22.1% (19.7-24.6) | 690 | 28.6% (25.4-32) | 276 | 23.2% (19.3-27.4) | 414 | 32.9% (27.3-38.8) |
| S positive / N negative | 205 | 7.6% (6.1-9.3) | 888 | 43.7% (40.1-47.4) | 692 | 74.3% (70.0-78.4) | 196 | 19.6% (15-24.8) |
| S negative / N positive | 92 | 3.0% (2.1-4.2) | 76 | 2.8% (1.8-4.3) | 0 | 0.0% (0.0-0.8) | 76 | 5.1% (2.8-8.4) |
| Unweighted total | 2706 |  | 2274 |  | 1003 |  | 1271 |  |

\*n denotes the number of samples tested
\*\* weighted by age, sex, ethnicity and sampling design

**Table 3.** **Unweighted and weighted** regional SARS-CoV-2 antibody positivity in unvaccinated primary school children and by COVID-19 vaccination status in secondary school children during Round 1 (10 November – 10 December 2021) in the SIS2 study**

| Region | Primary Total |  | Secondary Total |  | Secondary Unvaccinated |  | Secondary Vaccinated |  |
| --- | --- | --- | --- | --- | --- | --- | --- | --- |
|  | n* | Antibody positive (95CIs) | n* | Antibody positive (95CIs) | n* | Antibody positive (95CIs) | n* | Antibody positive (95CIs) |
| East Midlands | 225 | 39.5% (30.1-49.5) | 161 | 72.4% (62.3-81.1) | 110 | 46.7% (34-59.8) | 51 | 25.7% (12.5-43.2) |
| East of England | 382 | 50.3% (40.6-59.9) | 181 | 82.1% (73.8-88.7) | 101 | 32% (21-44.5) | 80 | 50.2% (35.6-64.8) |
| London | 366 | 34.8% (27.9-42.1) | 145 | 88.7% (78.1-95.3) | 66 | 38.5% (21-58.5) | 79 | 50.2% (32.7-67.7) |
| North East | 383 | 46.5% (40.7-52.4) | 354 | 75.1% (69.3-80.3) | 195 | 30.6% (22.9-39.2) | 159 | 44.5% (35.4-53.8) |
| North West | 418 | 49.3% (43.7-54.8) | 248 | 87.5% (81.7-91.9) | 126 | 41.7% (31.3-52.7) | 122 | 45.8% (35.4-56.5) |
| South East | 297 | 30.2% (22.6-38.8) | 239 | 79.7% (72.6-85.6) | 122 | 25.9% (16.7-37.1) | 117 | 53.7% (42.4-64.8) |
| South West | 254 | 21.9% (13.9-31.8) | 453 | 70.2% (64.7-75.3) | 262 | 35.6% (28.5-43.2) | 191 | 34.6% (26.6-43.3) |
| West Midlands | 134 | 53.8% (43.1-64.3) | 67 | 88.3% (70.4-97.3) | 28 | 40.1% (13.9-71.4) | 39 | 48.2% (24.2-72.8) |
| Yorkshire and the Humber | 247 | 39.1% (31.7-46.8) | 426 | 89.0% (83.6-93.1) | 261 | 68.5% (59.8-76.4) | 165 | 20.5% (11.7-31.9) |
| Total | 2706 |  | 2274 |  | 1003 |  | 1271 |  |

\*n denotes the number of samples tested
\*\* weighted by age, sex, ethnicity and sampling design

### Secondary school-aged children

The overall antibody positivity rate in secondary school students was 72.7% (1654/2274) and includes 44.1% (1003/2274) who had received ≥1 COVID-19 vaccine dose. After weighting and adjusting for test sensitivity, the national prevalence of SARS-CoV-2 antibodies was 82.4% (95%CI; 79.5-85.1), including 57.5% (95%CI; 53.9-61.1) in unvaccinated and 97.5% (95%CI; 96.1-98.5) in vaccinated students (**Table 2**). Antibody prevalence varied by region in unvaccinated and vaccinated students. In unvaccinated students, antibody prevalence ranged from 25.9% (95%CI; 16.7-37.1) in the South East to 68.5% (95%CI; 59.8-76.4) in Yorkshire and the Humber region while, in vaccinated students, antibody prevalence ranged from 20.5% (95%CI; 11.7-31.9) in Yorkshire and the Humber region to 53.7% (95%CI; 42.4-64.8) in the South East (**Table 3**). Antibody prevalence increased with age (p<0.001) (**Supplement Figure 2**), was not significantly different in urban versus rural students (p=0.1) and did not differ by sex (p=0.78).

### Comparison with seroprevalence in young adults

Adults aged 17-30 years, who had been eligible for COVID-19 vaccination earlier, had higher vaccine uptake for both doses than secondary school students with higher S-antibodies, through a combination of natural infection and vaccination, with non-overlapping 95%CI compared to secondary school students in four regions (East Midlands, North East, South East, and South West) (**Figure 2**). Primary school students were not vaccinated and had lower antibody levels in all English regions compared to secondary school students and young adults.

**Figure 2.**
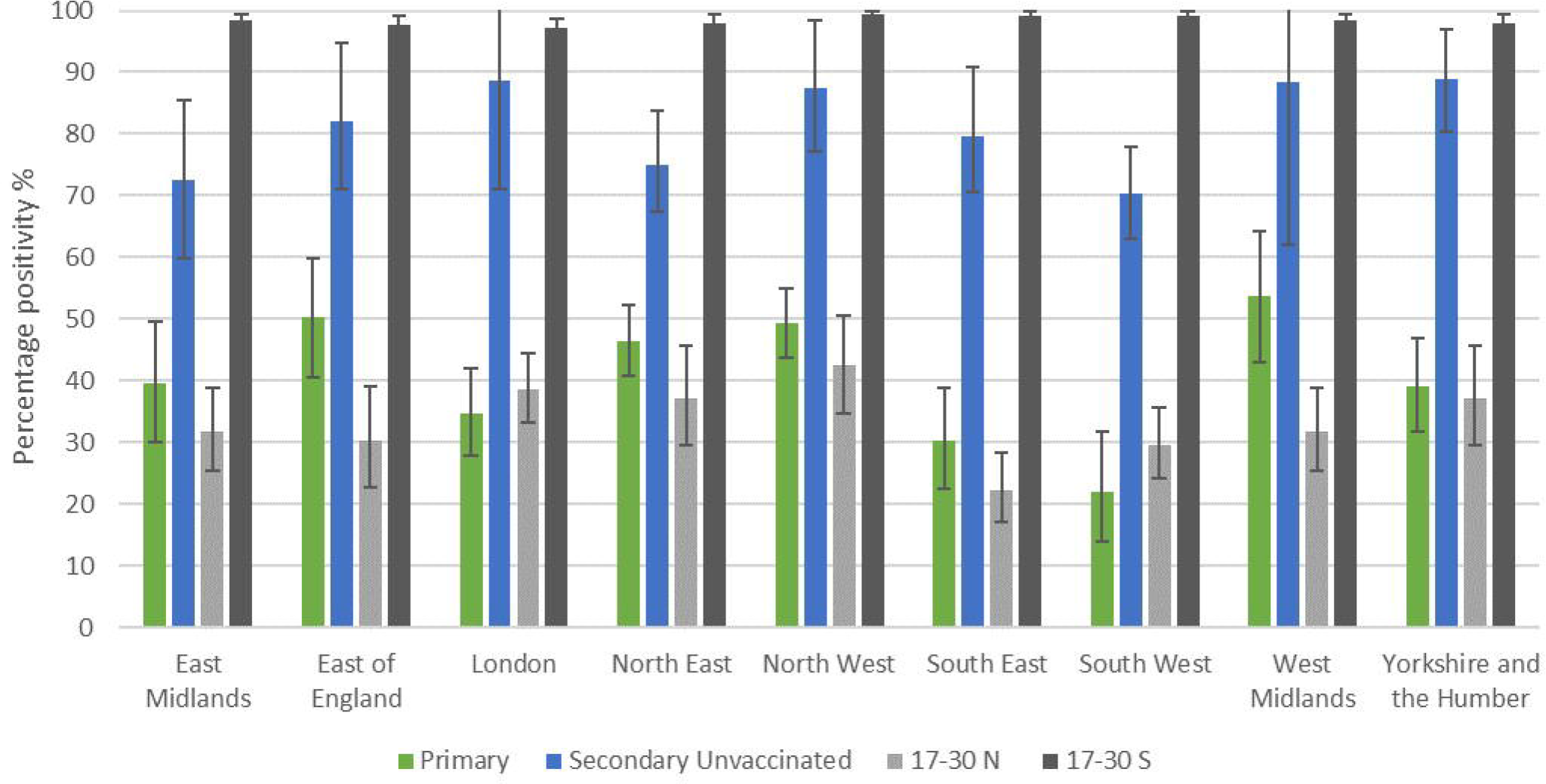
Regional weighted, adjusted total antibody prevalence in primary and secondary students participating in Round 1 (10 November – 10 December 2021) of SIS2 and N and S antibody positivity in 17-30-year-olds in NHSBT serosurveillance data. NB: NHSBT data uses NHS regions, therefore data for the Midlands was used for comparison with our cohorts in the East and West Midlands. North East data was used for comparison with our cohort in Yorkshire and the Humber. NHSBT data is weighted by age and sex.

## Discussion

We used an in-house OF assay developed and validated by UKHSA to estimate for the first time regional and national SARS-CoV-2 antibody prevalence after natural infection and vaccination in primary and secondary school-aged children in England.(13) In November 2021, an estimated 40.1% of primary school-aged students had infection-induced antibodies, while 82.4% of secondary school-aged children had SARS-CoV-2 antibodies through a combination of infection and vaccination, with 44% of our secondary school students having received ≥1 dose of COVID-19 mRNA vaccine ≥14 days prior to their antibody test. Across all regions, young adults, who became eligible for COVID-19 vaccination earlier than adolescents, had higher S-antibody prevalence than the students. These findings provide a baseline for monitoring national and regional trends in natural and vaccine-induced antibodies with increasing vaccine uptake and emergence of new variants in children and adolescents.

OFs have multiple advantages over serum antibody testing because they are easy to use on a large-scale, are non-invasive, painless, more acceptable to participants and can be used in challenging populations, such as young children, thereby allowing higher recruitment rates across geographical areas because specialist expertise is not needed to collect the sample when compared to blood sampling.(13) Although the sensitivity of our OF assay is lower than serum antibody assays, population estimates can be adjusted for its sensitivity and specificity. Adjustment was, however, not required for the vaccinated cohort because of higher sensitivity and specificity of the OF assay compared to serum (90-95%) achieved through the high antibody levels after vaccination, even with a single dose.

Our study provides the first snapshot of national and regional seroprevalence of SARS-CoV-2 antibodies in English children, when the first cohort of 12-15-year-olds became eligible for COVID-19 vaccination and highlights the importance of seroprevalence studies to assess population immunity and trends over time. Because of difficulties in sampling large numbers of children, there are few other large-scale, childhood national seroprevalence studies for comparison. In England, comparison with regional NHS-BT seroprevalence data in adults shows that SARS-CoV-2 antibody prevalence in children cannot be inferred from adult studies. By week 42 (beginning 17 October) 2022, COVID-19 vaccine uptake in young adults was 63-68%, for the first dose and 55-57% for the second dose.(17) Consequently, S-antibody prevalence rates were higher in young adults compared to secondary or primary school children. There were, however, wide regional differences in infection-induced and vaccine-induced SARS-CoV-2 antibody prevalence rates between these three cohorts, which will be further confounded with the on-going rollout of the childhood COVID-19 vaccine programme and emergence of new variants. Moreover, since most children have asymptomatic or mild, transient infection, they may not get tested for the virus and, therefore, extrapolating from community infection rates will likely grossly underestimate population prevalence of SARS-Cov-2 antibodies,(18) as evidenced by the substantially lower confirmed infection rates in our cohort.

In England, the ONS COVID-19 Infection Survey (CIS) recently extended its national seroprevalence surveillance using finger prick tests to measure serum antibodies in adults to include a small cohort of children as young as 8 years. Sampling between 29 November 2021 and 5 December 2021 estimated 33% antibody seropositivity for 8-11-year-olds and 81% for 12-15-year-olds, which is consistent with our national surveillance estimates. However, the small sample size of children in ONS SIS precluded regional estimates and a high threshold used to confirm SARS-CoV-2 antibody positivity may underestimate seroprevalence rates in children.(19) The availability of regional data on infection- and vaccine-induced antibody prevalence may be particularly useful for immunisers and policy makers for targeting vaccine uptake in more susceptible regions and populations across England.

In England and elsewhere, opportunistic testing of residual sera from children having blood tests, in hospital, for example, has been used to assess SARS-CoV-2 antibody prevalence in children but these are prone to multiple biases, especially representativeness to the general population.(20-24)

Internationally, one Mexican study did include a nationally representative sample of children along with adults to estimate an overall SARS-CoV-2 antibody seroprevalence of 24.9% during August-November 2020, including 22.5% in 1,891 children ;notably, two-thirds of all seropositive participants reported being asymptomatic with their infection.(25) In Portugal, national seroprevalence using 8,463 participants was estimated to be 15.5% across all age-groups during February-March 2021, including 14.3% among 721 children aged 1-9 years and 12.9% among 955 10-19 year-olds.(18)

### Interpretation of findings

The UK strategy of closing schools last and opening schools first compared to other settings meant that children continued to benefit from in-person schools throughout most of the pandemic, with closures only during national lockdown periods in March-May 2020 (first pandemic wave) and January-February 2021 (alpha variant wave). Confirmed cases in school-aged children generally fluctuated with community infection rates.(26) As more adults were vaccinated as part of the national rollout, however, childhood infection rates became more disconnected from adult infection rates, especially after all mitigations were removed from schools since the start of the 2021/22 academic year (September 2021).(12)

Contrary to other countries, the UK adopted a cautious approach to vaccinating adolescents and younger children because of their low risk of severe COVID-19 and initial concerns about vaccine-induced myocarditis after the second mRNA vaccine dose in young males.(5) Consequently, roll-out of the Pfizer-BioNTech mRNA vaccine in adolescents (the only vaccine authorised in this age-group) has been slow and, after a rapid early uptake through a school-based programme for 12-15 year-olds, vaccine uptake reached 32% for one dose on 01 November 2021 and then plateaued. A second dose for 12-15-year-olds is recommended with an interval of 12 weeks between doses. The vaccine has been shown to protect against the low risk of severe COVID-19 in adolescents,(27) but provides limited short-term protection against infection, transmission or mild disease, especially with the Omicron variant, which can evade both natural and vaccine-induced immunity.(28, 29) In 5-11-year-olds, a lower-dose (10 vs 30 μg) Pfizer-BioNTech mRNA vaccine was offered to high-risk children from February 2022, with extension to all 5-11 year-olds from April 2022.(7) The impact of vaccination will require close monitoring given that, unlike other countries, most 5-11 year-olds will have already been naturally exposed to SARS-CoV-2, especially after the large case numbers observed with the Omicron wave during December-March 2022.(30)

Infections with the Omicron variant have been observed in all age-groups, with high rates of primary infections as well as recurrent infections among both previously-infected and vaccinated individuals, but hospitalisations and fatalities have remained very low.(31) When children returned to in-person schooling in January 2022, cases increased in both secondary and primary school-aged students which, along with ongoing rollout of the childhood COVID-19 vaccination programme, will result in higher antibody prevalence rates in the coming months.(30)

### Strengths & Limitations

The strength of this study lies in the large, representative cohort that allowed regional and national estimation of SARS-CoV-2 antibodies in primary and secondary school children across England. The establishment of this cohort has allowed other assessments, including vaccine sentiment, long-COVID and wellbeing surveys, while longitudinal follow-up will allow monitoring of changes over time, especially given the on-going vaccination programme and emergence of new variants.

There are some limitations. We were unable to recruit the required 180 schools and student participant rates were lower than the predicted 25% in primary and 15% in secondary schools, which meant that we did not recruit the anticipated 16,000 students for Round 1, although we were able to estimate regional and national prevalence with adequate precision with the sample sizes achieved. On-going recruitment of schools and students means that we will have higher participation rates in further rounds. Another limitation is the waning of SARS-CoV-2 antibodies over time, especially N-antibodies which decline quicker than S-antibodies after natural infection.(3) This may underestimate SARS-CoV-2 antibody prevalence in our cohort because some children may have been infected up to 18 months prior to their antibody test, particularly because OF antibody levels may be 10- to 100-fold lower than in serum. We partly mitigated this limitation by measuring both N- and S-antibodies (which persists longer after natural infection) in all participants. The high S-antibodies achieved after even a single vaccine dose substantially increases the sensitivity of the OF assay (unpublished data). Unfortunately, there is no clear correlation between antibody positivity status or antibody thresholds and the risk of re-infection, especially with new variants that can evade prior immunity. Also, we have only presented qualitative results, further work is required to assess the use quantitative antibody levels with the OF assay to understand antibody protection against infection and waning over time. Finally, we compared our cohort with data from NHS-BT, with under-representation of minority ethnic groups, and may therefore not be representative of the young adult population nationally.

In conclusion, we developed and implemented a novel, non-invasive OF test on a large-scale to estimate national and regional SARS-CoV-2 antibody prevalence in children across England. In November 2021, 40.1% of primary school children and 82.4% of secondary school children had antibodies through natural infection and/or vaccination in England. On-going surveillance will be important to monitor the effects of vaccination, the Omicron variant and future variants in children.

## Ethical approval

This study was granted approval by UKHSA Research Ethics and Governance Group on 2/11/2021, R&D ref: 474

## Data Availability

De-identified study data are available for access by accredited researchers in the ONS Secure Research Service (SRS) for accredited research purposes under part 5, chapter 5 of the Digital Economy Act 2017. For further information about accreditation, contact or visit the SRS website.

