## Supplementary Material for "National and regional prevalence of SARS-CoV-2 antibodies in primary and secondary school children in England: the School Infection Survey, a national open cohort study, November 2021"

**SUPPLEMENT**

**Supplement 1. Number of schools in SIS1 and additional schools required to achieve a representative sample of schools for each region in SIS2**

| Region | Primary | | Secondary | |
| --- | --- | --- | --- | --- |
|  | Current sample | Additional required | Current sample | Additional required |
| East Midlands | 2 | 11 | 1 | 6 |
| East of England | 3 | 10 | 3 | 4 |
| London | 0 | 13 | 2 | 5 |
| North East | 13 | 0 | 7 | 0 |
| North West | 7 | 6 | 7 | 0 |
| South East | 3 | 10 | 3 | 4 |
| South West | 3 | 10 | 3 | 4 |
| Yorkshire and The Humber |  | 13 | - | 7 |
| West Midlands |  | 13 | - | 7 |
| **Grand Total** | **31** | **86** | **26** | **37** |

*Note: A sample of this size is only required to produce estimates of precision when anti-body prevalence in the school pupil population is close to 50%. We have simulated other prevalence scenarios (both higher and lower), where smaller sample sizes will enable us to achieve the equivalent levels of precision. If we sample this many schools we would expect to achieve approximately 20,000 participants / Antibody tests*

**Supplement Figure 1. Weekly incidence of SARS-CoV-2 cases per 100,000b of population from Week 30 2020 in primary and secondary school-aged children using national testing data for England.**

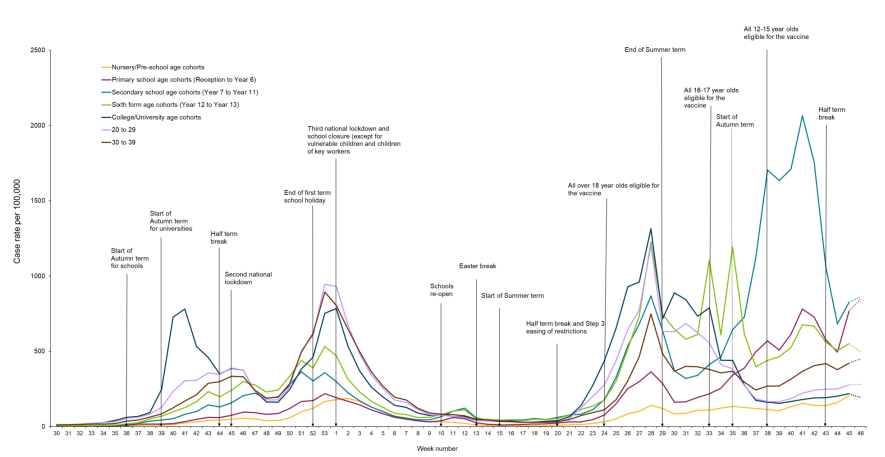

Graph taken from UKHSA Weekly Influenza and COVID-19 Surveillance graphs ^1^

Incidence definition: School age cohorts are calculated based on academic year birth cohorts. Those born between 01/09/2004 – 31/08/2005 are included in the year 12 school group and those born between 01/09/2003 – 31/08/2004 are included in the year 13 school group. Case rate denominators are sourced from ONS 2020 mid-year estimates.

**Supplement Figure 2. Weighted and adjusted antibody prevalence by age and vaccine status in SIS2 participants in Round 1 (10 November – 10 December 2021)**

Percentage positive (%)

| Age (years) | Unvaccinated | Vaccinated |
| --- | --- | --- |
| 4 | 31.61 | 0 |
| 5 | 30.99 | 0 |
| 6 | 34.82 | 0 |
| 7 | 38.05 | 0 |
| 8 | 42.72 | 0 |
| 9 | 44.93 | 0 |
| 10 | 57.93 | 0 |
| 11 | 50.64 | 6.11 |
| 12 | 33.15 | 46.21 |
| 13 | 37.89 | 48.42 |
| 14 | 28.9 | 57.82 |
| 15 | 48.66 | 48.39 |
| 16 | 33.18 | 65.57 |
| 17 | 48.88 | 47.34 |

1.	UKHSA. Weekly Influenza and COVID-19 Surveillance graphs. 2021. <https://assets.publishing.service.gov.uk/government/uploads/system/uploads/attachment_data/file/1035958/Weekly_COVID-19_and_Influenza_Surveillance_Graphs_w47.pdf> (accessed 12th December 2021).
